# Drug target Mendelian randomization supports apolipoprotein C3-lowering for lipoprotein-lipid levels reductions and cardiovascular diseases prevention

**DOI:** 10.1101/2023.11.04.23298088

**Authors:** Eloi Gagnon, Benoit J. Arsenault

## Abstract

**Background and aims:** Inhibitors of apolipoprotein C-III (apoC3) are currently approved for the reduction of triglyceride levels in patients with Familial Chylomicronemia Syndrome. We used drug target Mendelian randomization (MR) to assess the effect of genetically predicted decrease in APOC3 blood protein levels on cardiometabolic traits and diseases.

**Methods:** We quantified lifelong reductions in APOC3 blood levels by selecting all genome wide significant and independent (*r*^*2*^<0.1) single nucleotide polymorphisms (SNPs) in the *APOC3* gene region ±1Mb, from three genome-wide association studies (GWAS) of apoC3 blood protein levels (deCODE, n = 35,378, Fenland, n = 10,708 and ARIC, n = 7,213). We included the largest GWASes on 18 cardiometabolica traits and 9 cardiometabolic diseases as study outcomes.

**Results:** A one standard deviation lowering in apoC3 blood protein levels was associated with lower triglycerides, apolipoprotein B, density lipoprotein cholesterol, alanine aminotransferase, and glomerular filtration rate as well as higher high-density lipoprotein cholesterol levels. APOC3 lowering was also associated with lower risk of acute pancreatitis (odds ratio [OR] = 0.91 95% CI=0.82 to 1.00), aortic stenosis (OR = 0.82 95% CI=0.73 to 0.93), and coronary artery disease (OR = 0.86 95% CI=0.80 to 0.93), and was associated with increased parental lifespan (0.06 95% CI=0.03 to 0.09 years). These results were concordant across robust MR methods, the three protein datasets and upon adjustment for APOA1, APOA4 and APOA5 using a multivariable MR framework.

**Conclusions:** These results provide evidence that apoC3 lowering could result in widespread benefits for cardiometabolic health and encourage the launch of trials on apoC3 inhibition for coronary artery disease prevention.

## Introduction

Apolipoprotein C-III (apoC3) is an established target for triglyceride lowering. ApoC3 reduction by the antisense oligonucleotide (ASO) volanesorsen resulted in 77% lower circulating triglyceride levels in individuals with the rare congenital syndrome Familial Hyperchylomicronemia Syndrome (FCS) (Witztum et al. 2019). Beyond triglyceride lowering, APOC3 could be implicated in the pathogenesis of other diseases (Giammanco et al. 2023), opening potential opportunity for extending APOC3-ASO’s indications for other more common conditions.

Mounting genetic evidence support a causal role of apoC3 inhibition in the prevention of cardiovascular diseases. *APOC3* heterozygous loss of function mutations carrier have on average 40% lower plasma triglyceride levels and a 46% lower coronary heart disease risk in the exome sequencing project (498 carriers vs. 110,472 non-carriers) (The TG and HDL Working Group of the Exome Sequencing Project 2014) and 36% lower risk of ischemic heart disease in the Copenhagen General Population Study (302 carriers, 75,423 non-carriers) (Jørgensen et al. 2014). Akin to studies on loss of function carriers, mendelian randomization (MR) studies leverage naturally occurring genetic variations to investigate the impact of perturbing drug targets on health outcomes (Gill and Burgess 2022). These genetic variations are randomly distributed during meiosis and remain constant throughout life, making MR results unlikely to be influenced by confounding factors or reverse causality. Drug target MR study designs may also help identify new indications for existing drugs, predict randomized clinical trial (RCT) outcomes, and identify ontarget adverse effects (Gill and Burgess 2022). MR studies on apoC3 have proven challenging as this protein is encoded by a complex genetic locus on chromosome 11 harboring other genes influencing lipoprotein metabolism such as *APOA1, APOA4*, and *APOA5*. However, recent advances in large-scale proteo-genetics can help isolate genetically predicted apoC3 blood concentrations from these other apolipoproteins opening the door to drug target MR studies on apoC3 genetic inhibition.

Here, we performed drug target MR analyses evaluating the effect of genetically predicted lower apoC3 protein levels on CAD as well as 8 other cardiometabolic diseases and 18 cardiometabolic traits. We show that genetically predicted apoC3 reduction is associated with a healthier metabolic profile as well as a lower risk of CAD, aortic stenosis and acute pancreatitis.

## Methods

### Genome-wide association studies on blood protein levels

The datasets used to derive the study exposures and outcomes are presented Supplementary Table 1. We included three distinct population-based cohorts for genetic-plasma protein levels associations: deCODE, Atherosclerosis Risk in Communities (ARIC), and Fenland (Ferkingstad et al. 2021; Zhang et al. 2022; Pietzner et al. 2021). 1) In the deCODE cohort, plasma protein levels were measured in 35,365 Icelanders. These measurements were adjusted for age and sex and the resulting residuals were inverse rank normal transform prior to GWAS. Genotyping was performed using Illumina SNP chips. 2) In the ARIC population-based cohort, protein plasma levels were measured in 7,213 European Americans. These measurements were adjusted in a linear regression model including PEER factors and the covariates sex, age, study site, and 10 ancestry-based principal components. Genotyping was performed using Illumina SNP chips. 3) In the Fenland population-based cohort, protein plasma levels were measured in 10,708 European-descent participants. These measurements were adjusted for age, sex, the first ten principal components of ancestry and the test sites. The resulting residuals were inverse rank normal transform prior to GWAS. Fenland participants were genotyped using three genotyping arrays: the Affymetrix UK 1149 Biobank Axiom array (OMICs, n=8994), Illumina Infinium Core Exome 24v1 (Core-Exome, 1150 n=1060) and Affymetrix SNP5.0 (GWAS, n=1402). Primary analyses were performed using the deCODE cohort because this study had the largest sample size.

### Cardiometabolic traits and outcomes genome-wide association studies

Relevant information on the GWAS summary statistics used throughout this study is presented in Supplementary Table 1. Parental lifespan, fasting glucose, fasting insulin, and glomerular filtration rate were reported in years, log(pmol/L), log(pmol/L) and log(eGFR) respectively. For better interpretability and comparability, we transformed these summary statistics to a one standard deviation scale using the sdY.est function in the coloc package (Wallace 2020). The other continuous variables were already inverse-rank normal transformed in the GWAS.

### Genetic variant selection and harmonisation

We identified common variants (minor allele frequency > 0.01) within a 1 Mb window around the gene region associated (p<5e-8) with lower blood protein levels in the ARIC (N= 7,213), deCODE (N= 35,365) or Fenland (N= 10,708) population-based cohort. Variants were then clumped respectively to the lowest p-value of any of these cohorts (linkage disequilibrium *r*^*2*^ <0.1 and window = 10 MB). When a SNP alter the structure of a protein, the statistical association between that SNP and protein levels may be biased, a phenomenon known as epitope binding artifact. To prevent epitope binding artifacts, these SNPs were annotated using the variant effect predictor (McLaren et al. 2016). We removed every SNPs tagged as being missense variant, stop gained, stop lost, start gained, start lost, or frameshift. The strength of every instrument was evaluated with the Cragg-Donald F-statistic (Stephen Burgess, Thompson, and CRP CHD Genetics Collaboration 2011). Variant harmonization was performed by aligning the betas of different studies on the same effect allele with the *TwoSampleMR* V.0.5.6 package (Hemani et al. 2018). When a particular exposure SNP was not present in the outcome dataset, we used proxy SNPs instead (R2>0.8). We used the LD matrix of the 1000 Genomes Project - European sample of the Utah residents from North and West Europe.

### Mendelian randomization analyses

For univariable primary MR analyses, we performed the inverse variance weighted (IVW) method with multiplicative random effects (Stephen Burgess, Foley, and Zuber 2018). MR must respect three core assumptions (relevance, independence and exclusion restriction) for valid causal inference. Failure to respect these assumptions can occur if the genetic instruments influence several traits on different causal pathways. This phenomenon, referred to as horizontal pleiotropy, can be balanced by applying robust MR methods (Slob and Burgess 2020). To verify if pleiotropy likely influenced the primary univariable MR results, we performed four different robust MR analyses: the MR Egger intercept test (Bowden, Davey Smith, and Burgess 2015), the contamination mixture (Stephen Burgess et al. 2020), the weighted median, and the MR-PRESSO (Verbanck et al. 2018), each making a different assumption about the underlying nature of the pleiotropy. Consistent estimates across methods provide further confirmation about the nature of the causal links. An intercept not significantly different from zero in the Egger test support that pleiotropy is balance (Bowden, Davey Smith, and Burgess 2015). Univariable MR analyses were performed using the *TwoSampleMR* V.0.5.6 package (Hemani et al. 2018).

### Multivariable Mendelian randomization analyses

For multivariable primary MR analysis, we conducted the IVW method (S. Burgess and Thompson 2015). As robust MVMR analyses, we used the multivariable MR-Egger (Rees, Wood, and Burgess 2017), the multivariable median method, and the multivariable MR-Lasso method (Grant and Burgess 2021). Akin to robust univariable MR analyses, each method makes different assumptions about the underlying nature of the pleiotropy so that consistent estimates give confidence in the robustness of the causal finding. Multivariable MR analyses were performed using the *MendelianRandomization* V.0.9.0 package (Yavorska and Burgess 2017). To estimate instrument strength given the other exposures included in the model, we calculated conditional F statistics (Sanderson, Spiller, and Bowden 2021).

## Results

We identified up to 15 genetic proxies for apoC3 levels, explaining 3% of the variance in APOC3 levels (Supplementary Table 2 and Supplementary Table 3). Lower genetically predicted APOC3 plasma levels were associated with lower triglyceride levels (effect size per 1 SD [standard deviation] lower apoC3 plasma levels = -0.77 95% CI=-0.99 to -0.55, p=6.4e-12). This result was directionally consistent and maintained statistical significance (p-value < 0.05) across all robust MR approaches (weighted median, weighted mode, contamination mixture, MR-PRESSO) (Supplementary Table 4) and Egger intercept did not differ from zero (Supplementary Table 5). This result was similar across the other study cohorts. Altogether, these results corroborate RCT findings with RNA interference therapy targeted at *APOC3* providing validation for the genetic instrument.

MR analyses were consistent with apoC3 lowering being associated with a more favorable cardiometabolic risk profile. Genetically instrumented predicted apoC3 lowering was associated with lower apoB levels (−0.20 95% CI=-0.27 to -0.12, p=1.3e-06), lower LDL cholesterol levels (−0.16 95% CI=-0.22 to -0.11, p=4.2e-09), lower aspartate amino transferase (−0.08 95% CI=-0.13 to -0.04, p=5.9e-05), lower alanine amino transferase (−0.04 95% CI=-0.06 to -0.02, p=2.6e-05) and higher HDL cholesterol levels (0.37 95% CI= 0.27 to 0.47, p=5.0e-14, Figure 1). There was robust evidence of a small negative effect on glomerular filtration rate (−0.02 95% CI=-0.03 to 0.00, p=6.1e-03). There was nominal evidence for an association with fasting glucose, but not reaching statistical significance in robust MR analyses.

**Figure 1.**
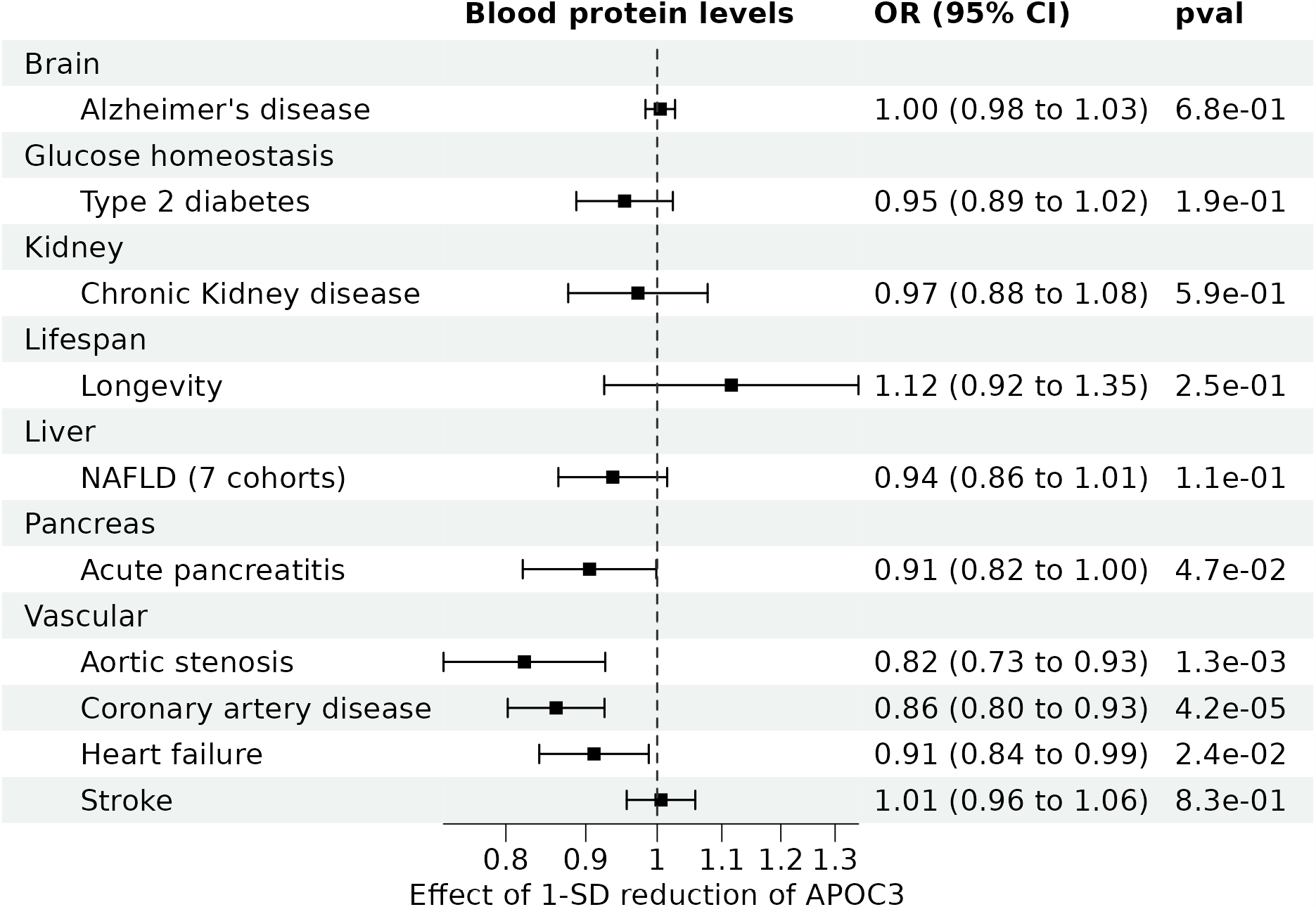
Effect of one standard deviation reduction in genetically predicted APOC3 blood protein levels on 18 cardiometabolic traits. Standard deviation change per one standard deviation increase with 95% confidence intervals are shown. P-values correspond to two-sided tests for the inverse variance weighted method with random effect model.

MR analyses were consistent with protective associations of genetically predicted apoC3 lowering with risk of aortic stenosis (OR per 1 SD reduction in APOC3 levels 0.82 95% CI=0.73 to 0.93, p=1.3e-03), and CAD (OR = 0.86 95% CI=0.80 to 0.93, p=4.2e-05, Figure 2). There was nominal evidence for an association with acute pancreatitis and heart failure but not reaching statistical significance in robust MR analyses. There was no evidence for an association with non-alcoholic fatty liver disease, type 2 diabetes, chronic kidney disease, Alzheimer’s disease, or stroke. There was evidence for an association with longer life expectancy (0.06 years 95% CI= 0.03 to 0.09, p=1.4e-05).

**Figure 2.**
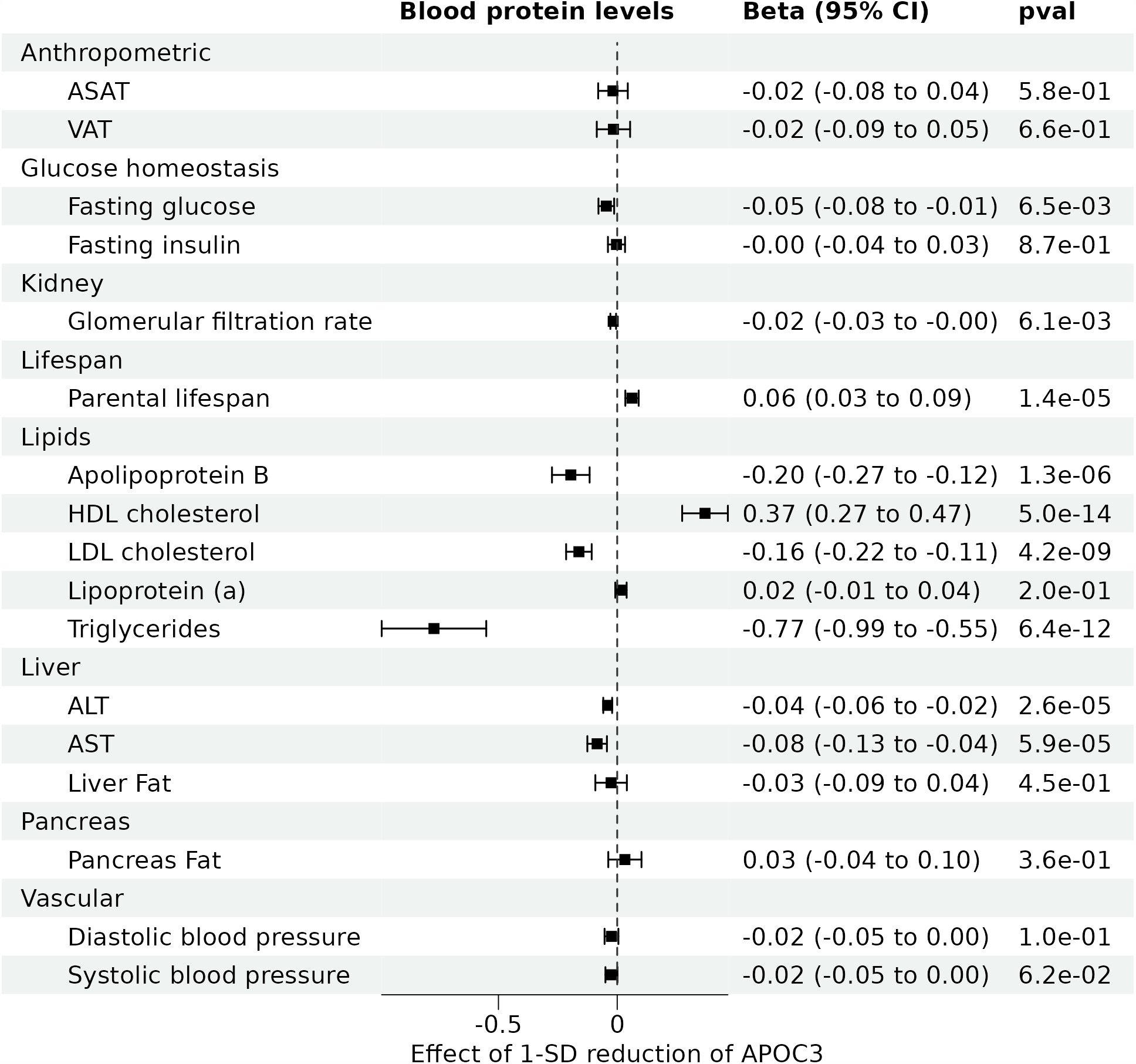
Effect of one standard deviation reduction in genetically predicted APOC3 blood protein levels on 9 cardiometabolic outcomes. Odds ratios per standard deviation with 95% confidence intervals are shown. P-values correspond to two-sided tests for the inverse variance weighted method with random effect model.

The human *APOC3* gene is located on chromosome 11 in a cluster with three other apolipoprotein genes, *APOA1, APOA4*, and *APOA5*. The genetic variants used to proxy apoC3 levels may also be associated with pleiotropic pathway through the levels of these apolipoproteins. We performed multivariable MR analyses, adjusting for genetic associations with apoA1, apoA4 and apoA5 plasma protein levels in the same cohort. We extracted the genetic instruments for these proteins using the same genetic instrument selection procedure as for *APOC3*. We then pooled these SNPs to the lowest p-value of any of the exposures, using the same parameter setting as the univariable MR (*r*^*2*^=0.1, window=1 Mb), resulting in 65 SNPs. Conditional F statistics were 20 for apoA1, 4 for apoA4, 89 for apoA5, and 5 for apoC3, indicating adequate instrument strength. Adjustment for genetic associations with serum apoA1, apoA4 and apoA5 did not influence the direction or the significance of the MR estimates (Supplementary Table 6). Adjustment for these apolipoproteins did not affect the results. For example, genetically predicted apoC3 lowering was associated with a 0.77 SD decrease in triglycerides without adjustment, and a similar 0.77 SD decrease in triglycerides with adjustment (Figure 3). ApoC3 was similarly associated to CAD with and without adjustment (Figure 4).

**Figure 3.**
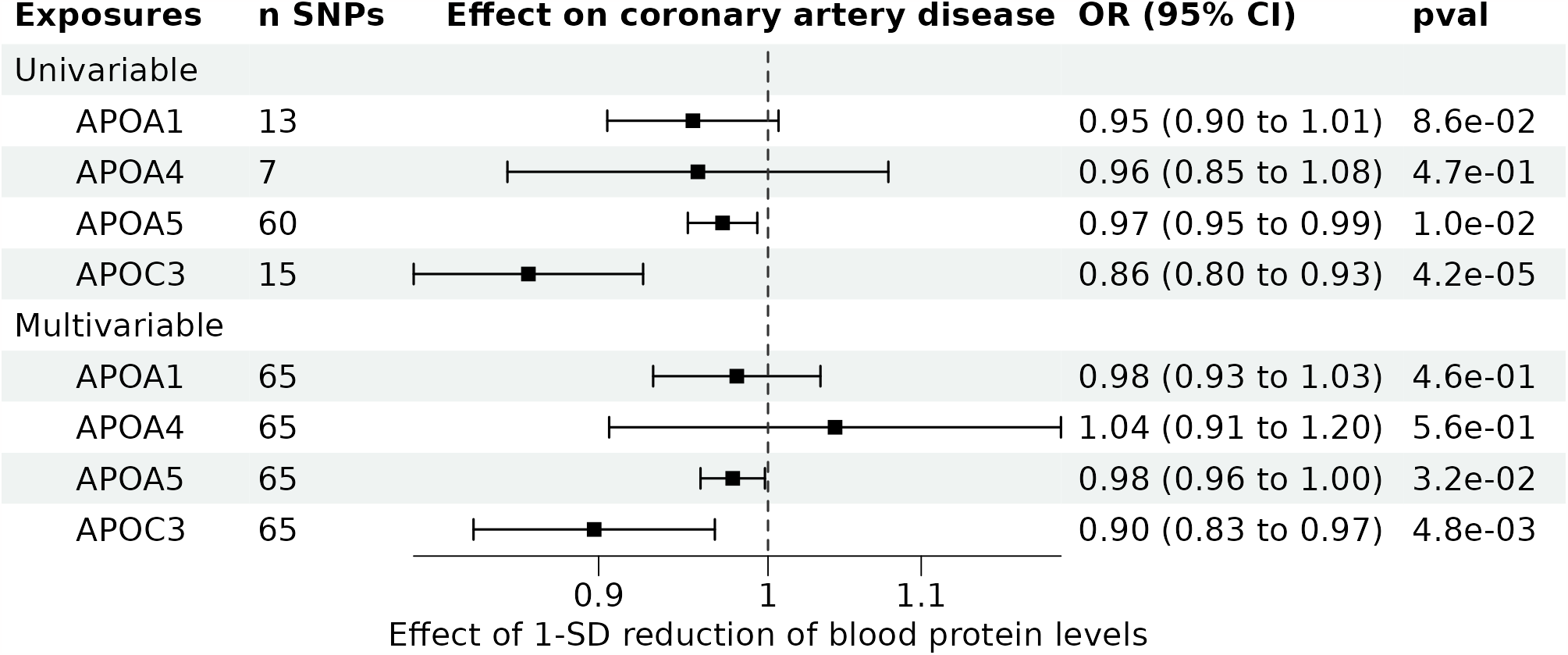
Effect of apolipoproteins in the APOA1/A4/A5/C3 gene cluster on triglycerides using univariable and multivariable Mendelian randomization. Standard deviation change per one standard deviation increase with 95% confidence intervals are shown. P-values correspond to two-sided tests. For univariable MR the IVW with random effect model was used. For multivariable MR, the multivariable IVW was used.

**Figure 4.**
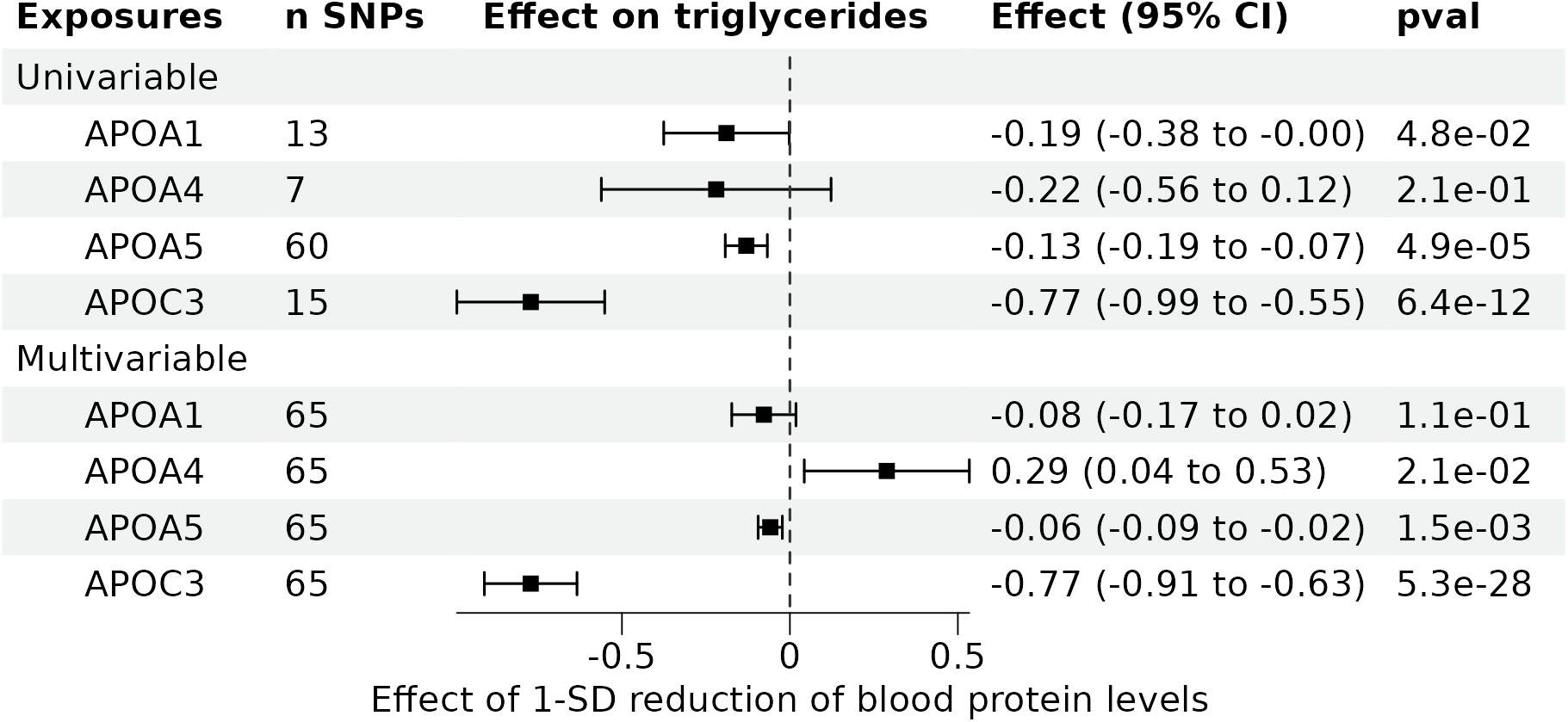
Effect of apolipoproteins in the APOA1/A4/A5/C3 gene cluster on coronary artery disease using univariable and multivariable Mendelian randomization. Odds ratios per standard deviation with 95% confidence intervals are shown. P-values correspond to two-sided tests. For univariable MR the IVW with random effect model was used. For multivariable MR, the multivariable IVW was used.

It is plausible that the effect of apoC3 lowering on the occurrence of diseases is linked to its influence on lipids. Specifically, triglycerides are a known causal factor of acute pancreatitis, while apoB is a known causal factor of coronary artery disease. To shed lights on the underlying mechanism, we used a multivariable MR approach (Figure 5, Supplementary Table 7). For this analysis, we included the genetic proxies of apoC3 identified in the univariable analysis as instrumental variables. When adjusting for triglyceride levels, apoC3 was no longer associated with acute pancreatitis. Similarly, when adjusting for apoB levels, the association between APOC3 and CAD was null. In contrast, when adjusting for either apoB or triglycerides, apoC3 remained associated with aortic stenosis, although the association did not reach statistical significance. These results support that the effect of apoC3 on CAD and acute pancreatitis is entirely explained by the effect on apoB and triglyceride levels. ApoC3 may have a direct effect on aortic stenosis, not mediated through its impact on lipoprotein-lipid levels.

**Figure 5.**
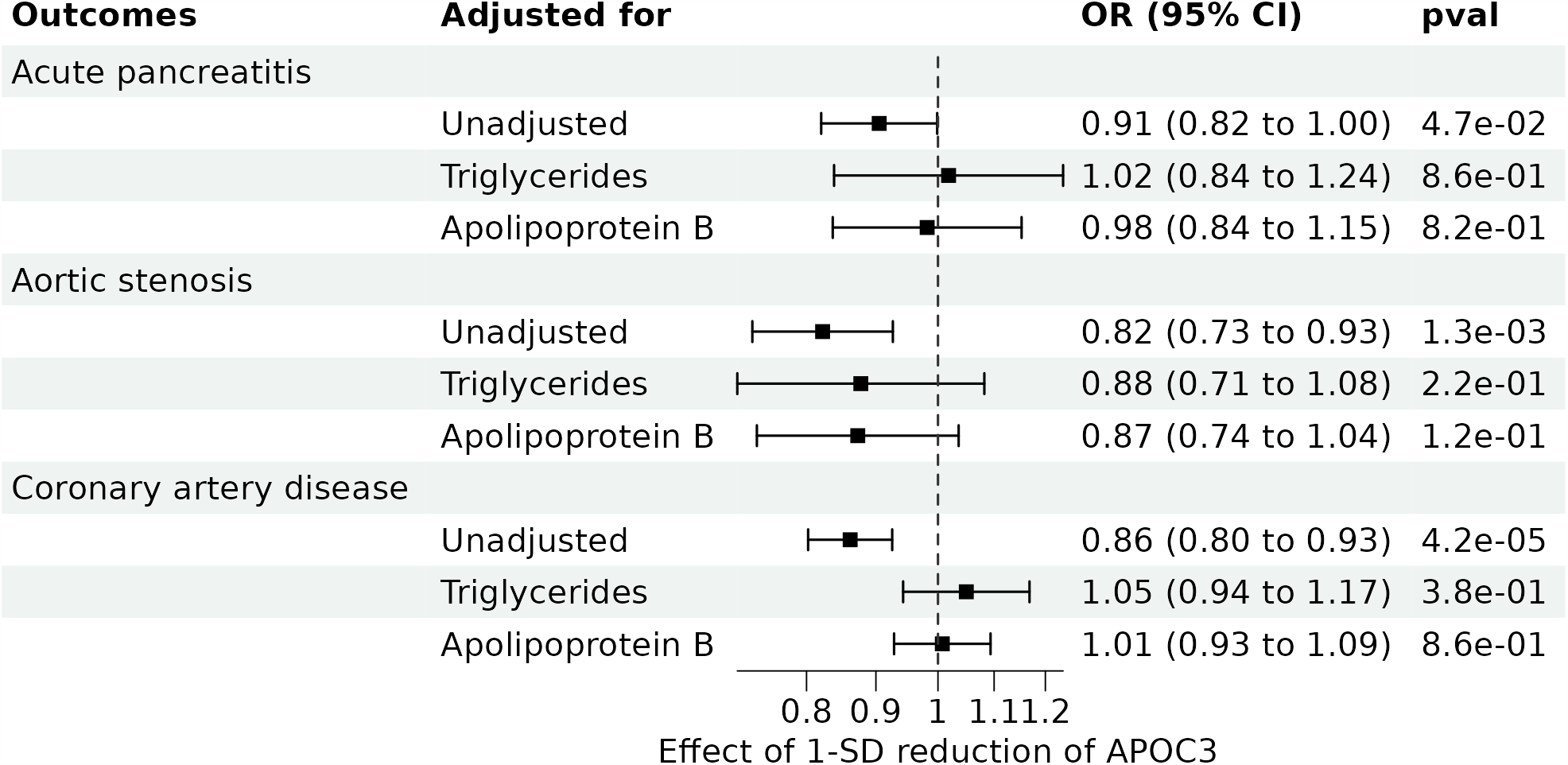
Effect of APOC3 on coronary artery disease, aortic stenosis, and acute pancreatitis before and after adjustment for triglycerides or apolipoprotein B levels using multivariable Mendelian randomization. Odds ratios per standard deviation with 95% confidence intervals are shown. P-values correspond to two-sided tests of the multivariable IVW analysis.

## Discussion

We performed drug target Mendelian randomization to determine the effect of genetically predicted apoC3 lowering on cardiometabolic health. Our results show that genetically predicted apoC3 lowering was associated with lower circulating levels of triglycerides, apoB, LDL cholesterol, alananine amino transferase, aspartate amino transferase, and higher circulating levels of HDL cholesterol. Furthermore, genetically predicted apoC3 lowering was associated with lower risk of CAD and aortic stenosis and to a smaller extent acute pancreatitis. These associations were concordant across three large blood pQTL datasets, concordant with robust MR methods and concordant in multivariable MR when including genetic information for apoA1, apoA4 and apoA5. Altogether, these results provide genetic evidence that apoC3 may represent a therapeutic target of interest to improve cardiometabolic health.

Most of our MR results are in line with results from RCTs on apoC3 lowering and extent those results to show that apoC3 reductions may provide cardiovascular benefits. A clinical trial involving patients with hypertriglyceridemia found that administering the ASO against *APOC3* olezarsen 50 mg every four weeks significantly lowered apoC3 by 74%, triglycerides by 23%, apoB by 10%, but no effect on lipoprotein(a) levels when compared to a placebo after 6 months of treatment (Tardif et al. 2022). Similarly, In a phase 1/2a study in healthy volunteers (ages 18– 65) without elevated triglyceride levels, administration of olezarsen every four weeks for three months resulted in a median reduction of circulating apoC3 of 89%, circulating triglycerides of 66%, circulating ApoB of 20%, and increase in HDL cholesterol of 60% observed immediately after the first injection and maintained throughout the treatment (Alexander et al. 2019). These concordant results provide external validation for our genetic instrument proxying lifelong *APOC3* perturbation.

The ASO against liver *APOC3*, volanorsen, is approved in the European Union for FCS (Paik and Duggan 2019). In this patient population, volanorsen reduced triglycerides by 77% when compared to placebo (Witztum et al. 2019). By doing so, Volanorsen could help reduce the rate of acute pancreatitis Observational data suggest that the incidence of acute pancreatitis increases approximately 3% for every increment of 100mg/dl in triglyceride levels over 1000 mg/dl (Rashid et al. 2016). In our MR study, genetically predicted *APOC3* lowering was associated with lower acute pancreatitis risk, although this assosication did not survive more robust MR analyses. Multivariable MR supported that this effect was presumably entirely explained by triglyceride levels. Although our MR analysis was not performed in the FCS population, these results provide genetic support to potenital benefits of apoC3 lowering in acute pancreatitis prevention.

Heterozygous carriers of rare loss of function mutation in *APOC3* were found to have 40% lower plasma triglyceride levels and a comparable lower CAD event rate (The TG and HDL Working Group of the Exome Sequencing Project 2014). Our findings indicate that CAD risk in the general population is influenced by apoC3 levels, not only apoC3 activity. Our multivariable MR results also suggest that the impact of apoC3 lowering is primarily driven by its influence on apoB. Whether apoC3 inhibitors could provide cardiovascular protection in patients with elevated triglyceride levels at high residual CVS risk will ultimately need to be determined in a large cardiovascular outcomes RCT.

Our MR results also suggest that apoC3 may have a role in the development of aortic stenosis. Aortic stenosis is characterised by thickening, fibrosis, and mineralization of the aortic valve leaflets (Moncla et al. 2023). Currently, there are no pharmacological therapy available to prevent or slow its progression. Surgical valve replacement remains the only treatment option. ApoC3 is present in aortic valve leaflets, as reported in a study by Capoulade et al. in 2020 (Capoulade et al. 2020). In this study, elevated levels of ApoC3-Lipoprotein(a) complexes have been shown to predict rapid hemodynamic progression of aortic stenosis in patients with mild to moderate aortic stenosis. High circulating levels of Lipoprotein(a) is an established causal factor for aortic stenosis (Arsenault et al. 2014; Thanassoulis et al. 2013). Our study results show that genetically predicted apoC3 lowering is associated with lower aortic stenosis risk but is not associated with lipoprotein(a) levels. These findings suggest that apoC3 may elevate the risk of aortic stenosis, but it does so independently of any impact on lipoprotein(a) levels. As an important proportion of the apoC3-aortic stenosis relation was mediated with lipoprotein-lipid levels, ApoC3 could represent an interesting therapeutic target for the prevention or treatment of aortic stenosis in patients with elevated triglyceride levels.

The main strength of the present study is the use of a MR study design using multiple robust and independent cis-acting instruments, minimizing the likelihood of horizontal pleiotropy. A second strength is the use of multiple study cohorts to derive the genetic proxies improving robustness. Our study findings must be evaluated within three main study limitations. First, genetic data was restricted for individuals of European ancestry to reduce chances of type 1 error due to population stratification, so the generalisability of the study’s findings to other populations is uncertain, especially in the context of other studies documenting a role of apoC3 in other ethnic groups (Saleheen et al. 2017). Second, MR evaluates lifelong effect of a genetically perturbed target, but trials often start later in life and for a shorter duration. Therefore, drug target MR typically estimates stronger causal effects compared to RCTs. Finally, we could only evaluated on-target effect of apoC3 lowering. Potential off-target effects of existing drug targeting APOC3 are not captured by our study design.

In conclusion, these results support that lowering of apoC3 blood protein levels may have several cardiometabolic health benefits such as a reduction in the risk of CAD, aortic stenosis and potentially acute pancreatitis and increased lifespan. RNA interference therapies effectively lowering apoC3 are already approved for the treatment of FCS. These findings provide genetic support and a rationale for the launch of cardiovascular outcomes RTCs on long-term apoC3 inhibition.

## Declarations

### Institutional Review Board Approval

All data used in this study are in the public domain. All participants provided informed consent and study protocols were approved by their respective local ethical committees. This project was approved by the Institutional Review Board of the Quebec Heart and Lung Institute.

### Data Availability

All data used in this study are in the public domain. Supplementary Table 1 describes the data used and relevant information to retrieve the summary statistics.

### Code Availability

Code to reproduce the results of this manuscript is available on https://github.com/gagelo01/APOC3

## Acknowledgements

We would like to thank all study participants as well as all investigators of the studies that were used throughout the course of this investigation.

## Authors’ contributions

Data acquisition and analysis EG. Conception and design EG, BJA. Drafting of the work EG, BJA. Both authors approved the final version of the manuscript.

